# Patient-Centered Approach Might Effectively Tackle The Definition Of Progression In Chronic Neurological Diseases: Results From The EmBioProMS Trial In Progressive Multiple Sclerosis

**DOI:** 10.1101/2021.09.07.21262777

**Authors:** Ahmed Abdelhak, Markus Krumbholz, Makbule Senel, Joachim Havla, Uwe K. Zettl, Ingo Kleiter, Thomas Skripuletz, Alexander Stahmann, Andre Huss, Kai Antweiler, Stefan Gingele, Markus C. Kowarik, Muna-Miriam Hoshi, Sandra Hengstebeck, Tim Friede, Albert C. Ludolph, Tania Kümpfel, Ulf Ziemann, Hayrettin Tumani

## Abstract

**Background:** Proper identification of disability accumulation in the routine clinical care of progressive multiple sclerosis (PMS) patients is usually a challenging task. Patient-reported outcome measurements (PROMs) can provide a practical, cost-efficient, and remotely accessible tool to assess disease progression.

**Methods:** EmBioProMS is a prospective, multicentric cohort, conducted in 7 specialized MS centers in Germany. PROMs were evaluated at inclusion and compared between patients with retrospective evidence of disease progression in the last two years and those with stable disease. Patients with either primary or secondary progressive MS according to the McDonald criteria 2017 were included in the analysis, while patients with incomplete PROMs scores, MS relapses, other neurological or systemic inflammatory diseases were excluded. The disease progression was assessed using a combined outcome parameter, including EDSS score, timed 25-foot walk test, and nine-hole-peg test.

**Results:** 185 patients were included in the final analysis (SPMS, n=77; PPMS, n=108). The median age and disease duration were 55 years and 13 years, respectively. Disease progression was diagnosed in 114 of 185 patients (61.6%). BDI-II, MSIS-29, and FSMC scores were worse in patients with evidence of disease progression in the last two years. Patients with any of the included PROMs above the 90th percentile had an odds ratio of 3.8 (95% confidence interval: 1.4–10.6, P=0.01) for having progression in the last two years in a binomial regression model adjusted for age, sex, disease duration, treatment status, center effect, and Expanded Disability Status Scale (EDSS). Similar results were observed in patients with PROM scores in the 80th and 70th percentile (OR: 2.9 and 3.7, P=0.015 and 0.003, respectively).

**Conclusion:** PROMs can be a simple and effective way to detect disability worsening in a chronic neurological disease like PMS and, therefore, substantially contribute to better classification and prognostication of the disease course through objective and structural patient-doctor communication.

**Trial Registration:** German Clinical Trials Register (Deutsches Register Klinischer Studien - DRKS), DRKS00020132

## Introduction

With the recent introduction of effective disease-modifying treatments (DMTs) for progressive multiple sclerosis (PMS), the accurate description and identification of disease progression have become more critical than ever. In routine clinical settings, disability worsening is mainly identified through detailed medical history and assessing the Expanded Disability Status Scale (EDSS) scores over longitudinal patient encounters. Nevertheless, several factors might hinder the accurate evaluation of the disability worsening. Clinical worsening is usually insidious and vaguely described by patients. Specifically, symptoms such as depression, fatigue, and cognitive deterioration are challenging to be objectively documented, liable to recall bias, and are poorly reflected in the EDSS score. Beyond that, longitudinal data of EDSS scores are not necessarily available. In addition, retrospective re-evaluation of EDSS scores based on the reported walking distance from patients could lead to inaccurate estimation of disability severity in up to 25% of the cases [1]. Although different ancillary investigations were proposed to detect progression, including various advanced MRI parameters, OCT scans, and body fluid biomarkers [2-6], none of the above-mentioned tools has been established in routine care, as longitudinal studies with large sample size are still required.

Over the last years, the assessment of patient-reported outcome measurements (PROMs) is getting into the focus of management of numerous chronic inflammatory conditions like rheumatoid arthritis and chronic inflammatory bowel disease (IBD) as recommended by many health authorities like the Food and Drug Administration (FDA). In IBD, PROMs could reflect the inflammatory disease activity, endoscopic results, and treatment success [7, 8]. In the field of neurology generally, and in MS specifically, PROMs are mainly applied to the settings of clinical trials [9], but not regularly used in the routine clinical care of people with MS (pwMS). MS-specific PROMs offer a standardized platform to assess symptoms such as fatigue, depression, and deterioration of the overall health [9].

In this work, we assessed a detailed battery of PROMs in relation to the evident clinical disease progression in a prospective multicentric cohort in Germany [10]. We aimed to identify clinically applicable cut-off values to differentiate between cases with disease progression and those with stable PMS. Our battery included three established scores in the MS care: Fatigue Score of Motor and Cognition (FSMC) [11], Beck Depression Inventory-II (BDI-II) [12], and Multiple Sclerosis Impact Scale-29 (MSIS-29) [13]. All three scores cover specific yet different MS symptoms, are characterized by high test-retest reliability, and can be performed in about 5 min each [9].

## Methods

### Study Design

EmBioProMS is a pilot, observational, prospective, multicentric study[10]. In summary, 218 patients with PMS according to the 2017 McDonald criteria and history of relapse-independent progression at any time were recruited in 7 centers in Germany. PPMS was defined according to the 2017 McDonald criteria [14], while SPMS was defined as patients with previous RRMS (fulfilling the 2017 McDonald criteria) who developed relapse-independent disability for at least one year. Patients with RRMS, with a history of relapse in the last three months, and other inflammatory or non-inflammatory diseases of the central nervous system, were excluded from our trial. Data regarding the disease onset, date, and symptoms of first manifestation, symptoms of the first manifestation, date of the diagnosis, number of documented relapses, duration of the progressive phase, number of documented relapses in the last two years, date of the most recent relapse, current, and previous DMT, and concomitant diseases were recorded at the baseline visit. At baseline medical history, EDSS, Nine-Hole Peg Test (9-HPT) and Timed 25-Foot Walk Test (T25FW) were evaluated through a certified EDSS rater. Patients were classified into two groups depending on treatment with DMT (treated vs. untreated). Patients who received the treatment up to the day of baseline visit or in a predefined period before the visit were included in the treated group, depending on the treatment (corticosteroids in the last 30 days; any interferon preparation, glatiramer acetate, natalizumab, dimethyl fumarate, teriflunomide, fingolimod, methotrexate, or azathioprine in the last three months; rituximab, ocrelizumab, or mitoxantrone in the previous 12 months; or cladribine or alemtuzumab in the previous 24 months). Patients who received the last dose beyond the specific period mentioned above for each medication or who were treatment-naïve at the baseline visit were assigned to the untreated group. Data collection and -management is conducted as a project within the platform of the German MS-Register by the German MS Society[15].

### Evaluation of the disease Progression

Determination of disease progression’s status (evidence of disease progression (EDP) vs. no evidence of disease progression (NEDP)) was determined based on the changes of EDSS, 9-HPT, and T25FW) compared to the previous study visit. The disease progression at the baseline visit was retrospectively assessed based on the documented clinical scores from the last two years. The disease course was considered “progressive” in the following cases: an increase in the EDSS score by 1 point in cases where the previous EDSS score was below 5.5, an increase in the EDSS score by ≥0.5 points in cases where the last EDSS score was ≥5.5, or an increase in the 9-HPT score or the T25FW score by 20% or more[16].

### Patients’ Reported Outcome Measurements (PROMs)

Three PROM questionnaires were collected at the baseline visit: FSMC, BDI-II, and MSIS-29. FSMC has 20 items, 10 of which focus on cognitive fatigue and 10 on motor fatigue. Each item is rated from one to five. The minimum value is 20 (no fatigue), and the maximum value is 100[11]. BDI-II is a 21-item self-report multiple-choice inventory. Each item is rated from zero to three, based on the severity of the answers. The minimum score is zero, and the maximum score is 63 points[12]. MSIS-29 consists of 29 items. Of them, 20 items constitute the physical scale, while nine questions form the psychological scale. Each item is rated from one to five according to the severity. Each scale is scored separately and converted to a 0–100 scale[13]. Only baseline visits with available scores from all three PROMs scales were included in this analysis.

### Statistical Analysis

Appropriate summary statistics were applied to describe the different variables, i.e., median with interquartile range for continuous variables and frequencies (percentages) for categorical variables. Nonparametric Spearman’s correlation coefficients were used to test the correlations between various parameters. The comparison for continuous variables between categorical groups was performed using the Mann–Whitney U test. Binomial logistic regression models and Linear models were applied to analyze the relationship between the PROMs scores, their percentiles, and progression status after adjusting for age, sex, disease duration, treatment status, baseline EDSS, disease phenotype (PP/SPMS), and center effect. Most models were built with evidence of disease progression as dependent variable to facilitate the clinical interpretation. Building the model using the disease progression as an independent variable resulted in similar results.

### Ethics approval

The study has been approved by the Institutional Review Board at the University Hospital of Ulm (#270/12), and the local ethical committees at the University Hospitals of Tuebingen, Ludwig-Maximilians University, Rostock, and Hanover. All patients signed informed consent before participating in the trial. Study procedures were conducted in accordance with the principles of the Declaration of Helsinki.

## Results

### Clinical Characteristics

Of the 218 patients, 33 were excluded due to incomplete PROMs or progression data, while 185 were included in the final analysis. A total of 77 (41.6%) patients had SPMS, while 108 (58.4%) had PPMS. The detailed clinical characteristics are summarized in Table 1.

**Table 1.** Clinical characteristics of the included patients.

| **Table 1.: Clinical characteristics of the included patients.** |  |  |  |  |
| --- | --- | --- | --- | --- |
| **Median (25–75 percentile), unless mentioned otherwise** | **SPMS (n=77)** | **PPMS (n=108)** | **Total (n=185)** | ***P*-value (SPMS vs. PPMS)** |
| **Age in years** | 55 (49–61) | 55 (50–60) | 55 (50–61) | n.s |
| **Disease duration in years** | 20 (15–30) | 9 (5–14) | 13 (7–23) | < 0.001 |
| **Female (%)** | 44 (57.1) | 58 (53.7) | 102 (55.1) | n.s |
| **Treated patients (%)** | 34 (44.2) | 48 (44.4) | 82 (44.3) | n.s |
| pulse steroid therapy (3-month interval) | 15 | 16 | 31 |  |
| Ocrelizumab | 2 | 27 | 29 |  |
| Interferon beta | 4 | 0 | 4 |  |
| Others | 13 | 5 | 18 |  |
| **EDSS score** | 6.0 (4.0–6.0) | 4.0 (3.5–6.0) | 4.5 (3.5–6.0) | < 0.001 |
| **Number of documented events of EDSS worsening (%)** | 35 (45.5) | 65 (60.2) | 100 (54.1) | n.s |
| **Number of documented events of T25FW worsening (%)** | 8 (10.4) | 17 (15.7) | 25 (13.5) | n.s |
| **Number of documented events of 9-HPT worsening (%)** | 10 (13.0) | 14 (13.0) | 24 (13.0) | n.s |
| **Number of documented events of progression of any kind (%)** | 43 (55.8) | 71 (65.7) | 114 (61.6) | n.s |
| SPMS, secondary progressive multiple sclerosis; PPMS, primary progressive multiple sclerosis; EDSS, Expanded Disability Score Scale; T25FW, Timed 25-Foot Walk Test; 9-HPT, Nine-Hole Peg Test. *P*-value for Mann–Whitney U test, n.s., non-significant |  |  |  |  |

Most of the disease progression events were confirmed through EDSS progression (*n*= 100, 87.7%). In 10.5% of the patients (*n*= 12), all three parameters (EDSS, T25-FW, and 9-HPT increase) for the definition of progression were fulfilled. As expected, SPMS patients have longer disease duration and higher EDSS scores (Table 1). 61 patients never received a treatment with DMT at the study inclusion (*n=* 13 and 48 with SPMS and PPMS, respectively). In the treated group (*n*= 82), the most common treatments administered at baseline were pulse steroid therapy every three months (*n*= 31) and ocrelizumab (*n*= 29). 4 patients received treatment with one of the interferon-beta medications at the time of evaluation of the PROMs.

### PROM Results

The scores of the included PROMs are summarized in Table 2. FSMC and MSIS-29 psychological (MSIS_psychological_ -29) component correlated inversely, but weakly, with age with a Spearman’s rho of -0.2 for all parameters (*P*= 0.016 and 0.027 respectively). Moreover, we found a moderate correlation between the EDSS with the MSIS_physical_ -29 and the FSMC (Spearman’s rho: 0.52 and 0.32, respectively, *P*< 0.001 for both). All included PROMs correlated moderately with the cerebral functional score (Spearman’s rho= 0.4 (*P<* 0.001) for FSMC, and 0.3 for the remaining PROMs (*P=* 0.001 for all)). None of the included PROMs correlated with the pyramidal functional score.

**Table 2.**
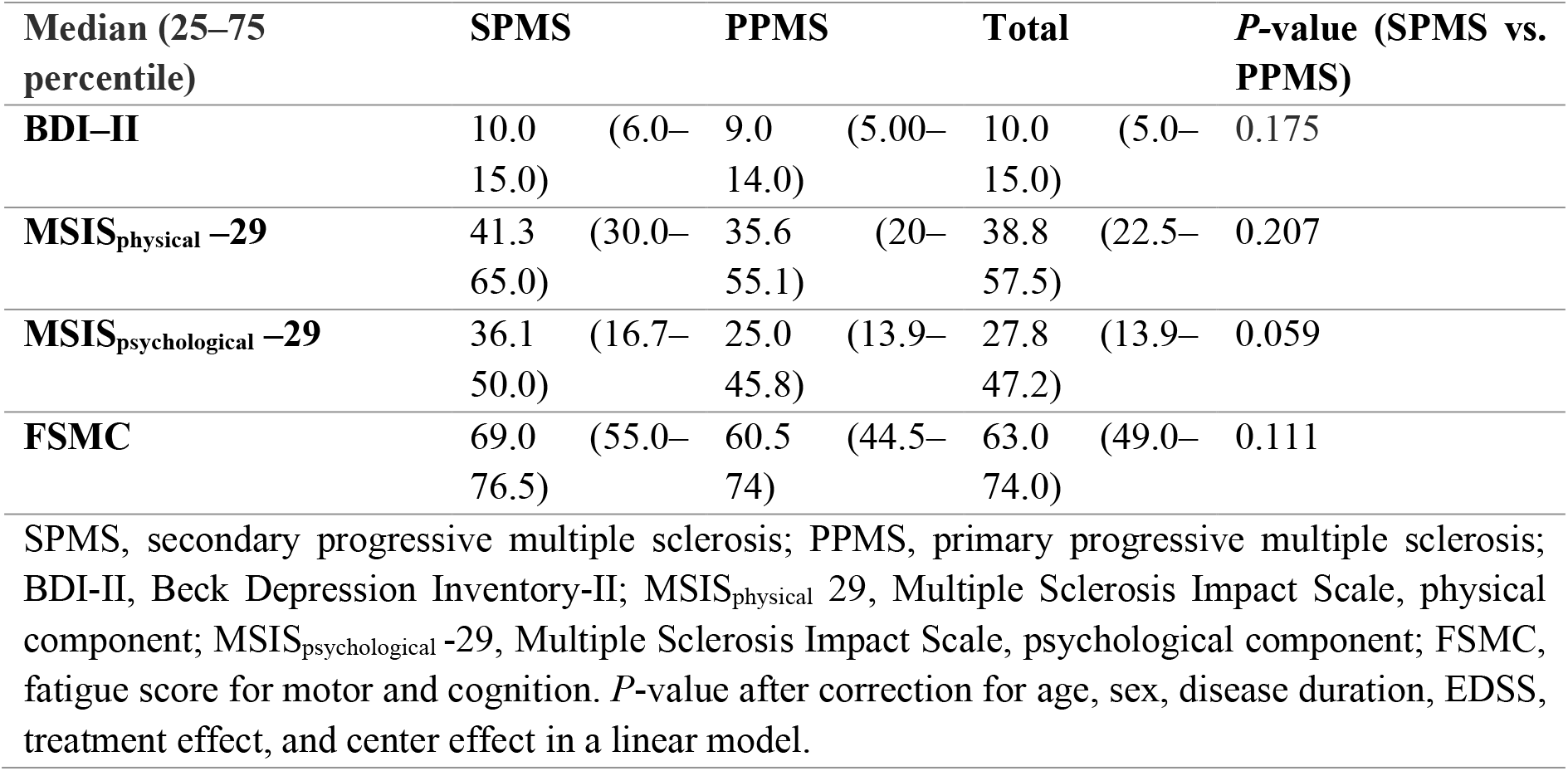
Patient-reported outcome measurement scores.

### PROMs in Correlation with Disability Progression

Patients with NEDP performed significantly better in all the tested PROMs than those with EDP, except in the FSMC score (Table 3, Fig. 1). Of the patients with any of the PROM scores above the 90^th^, 80^th^, and 70^th^ percentiles, 81.4% (35/43), 78.6% (55/70), and 74.5% (70/94), respectively, had EDP in the last two years.

**Table 3.**
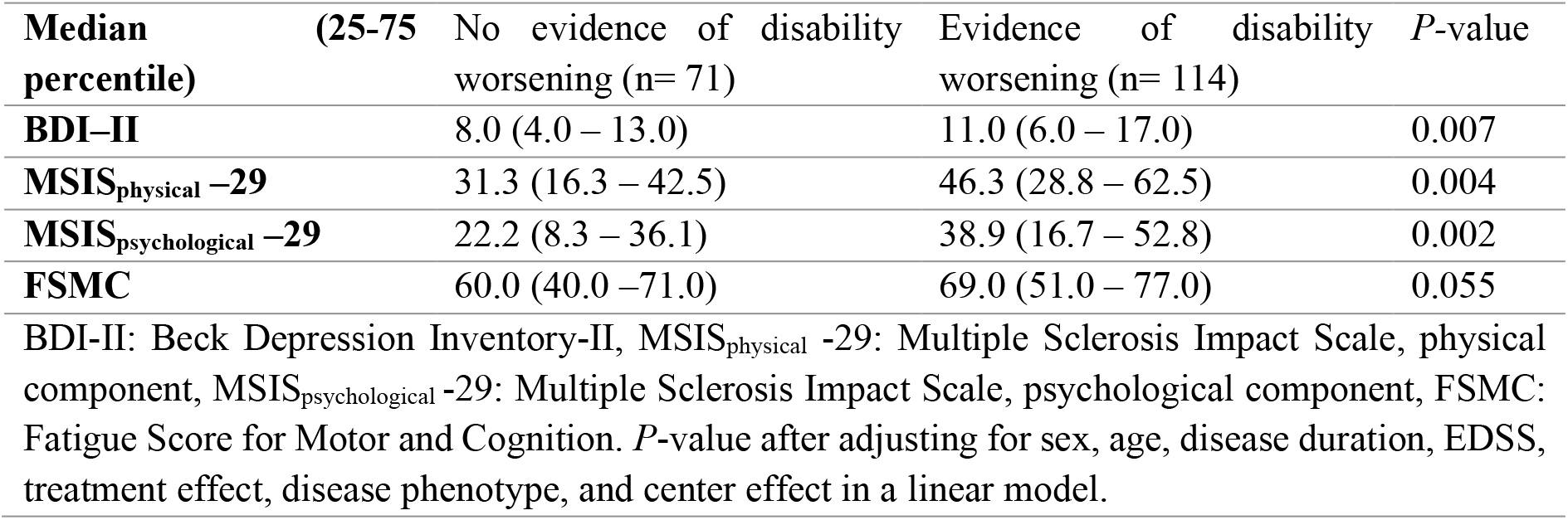
PROMs in patients with and without evidence of disease progression.

**Figure 1.**
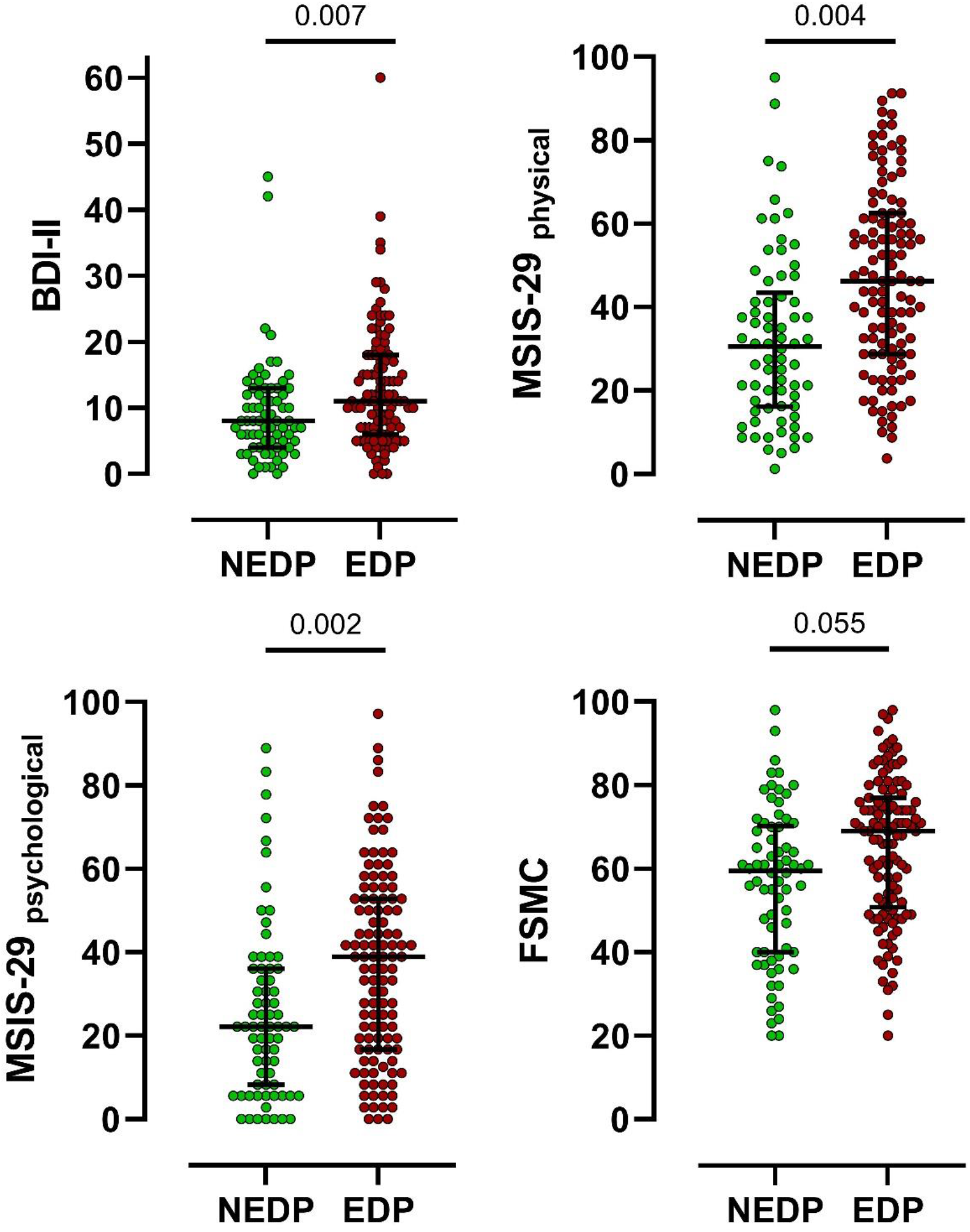
Patient-reported outcome measurements (PROMs) in cases with and without evidence of disability progression. BDI-II, Beck Depression Inventory-II; MSIS_physical_ -29, Multiple Sclerosis Impact Scale, physical component; MSIS_psychological_ -29, Multiple Sclerosis Impact Scale, psychological component; FSMC, fatigue score for motor and cognition. *P*-values after adjusting for sex, age, disease duration, EDSS, treatment effect, disease phenotype, and center effect in a linear regression model.

Applying a binomial logistic regression model adjusting for age, sex, disease duration, disease course, treatment status, center effect, and baseline EDSS score, patients with any of the PROMs scores above the 90^th^ percentile showed higher odds ratio (OR) for EDP in the last two years (OR: 3.8, 95 CI: 1.4– 10.6, *P*= 0.01, Fig. 2). Sensitivity analysis revealed a similar pattern at lower percentiles (e.g., 80^th^ and 70^th^) and for single PROMs scores separately (Table 4).

**Figure 2.**
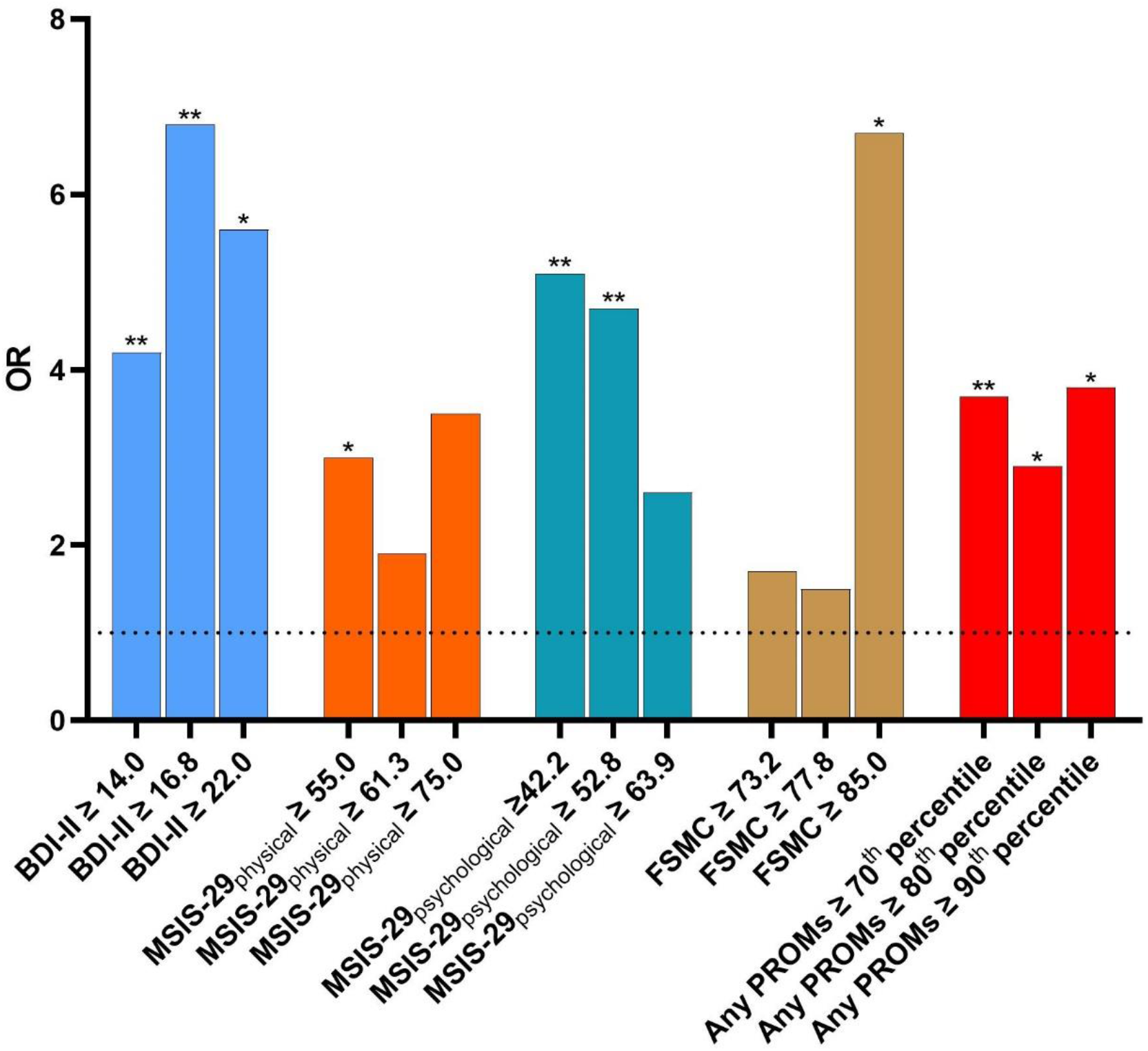
Distribution of the Odds Ratios (OR) for having disability progression in the preceding two years based on the selected PROMs cut-off values: BDI-II, Beck Depression Inventory-II; MSIS-29_physical_, Multiple Sclerosis Impact Scale, physical component; MSIS-29_psychological_, Multiple Sclerosis Impact Scale, psychological component; FSMC, fatigue score for motor and cognition. *:*P*<0.05, **:*P*<0.01. *P*-values after adjusting for sex, age, disease duration, EDSS, treatment effect, disease phenotype, and center effect in a binomial regression model.

**Table 4.**
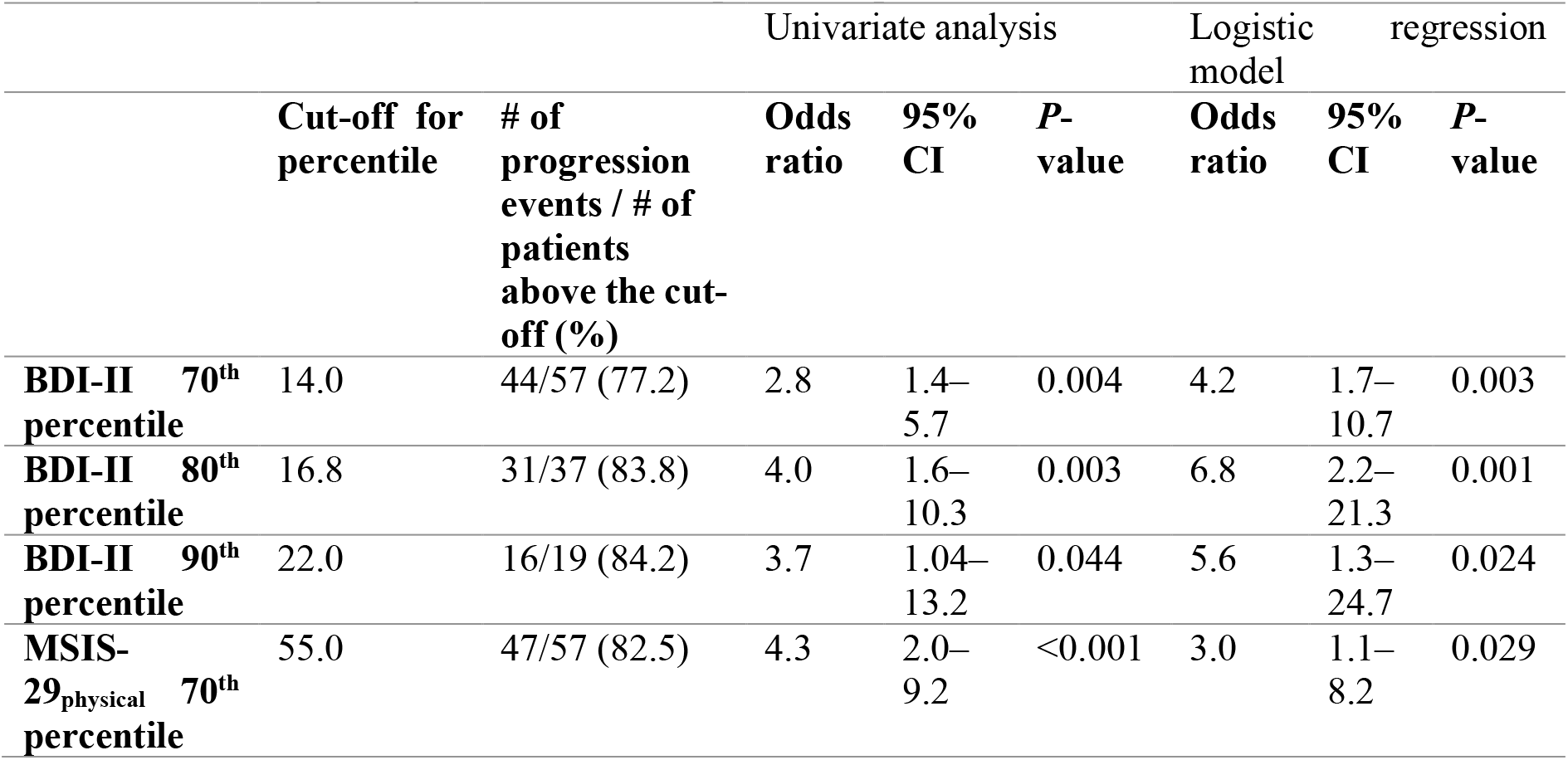
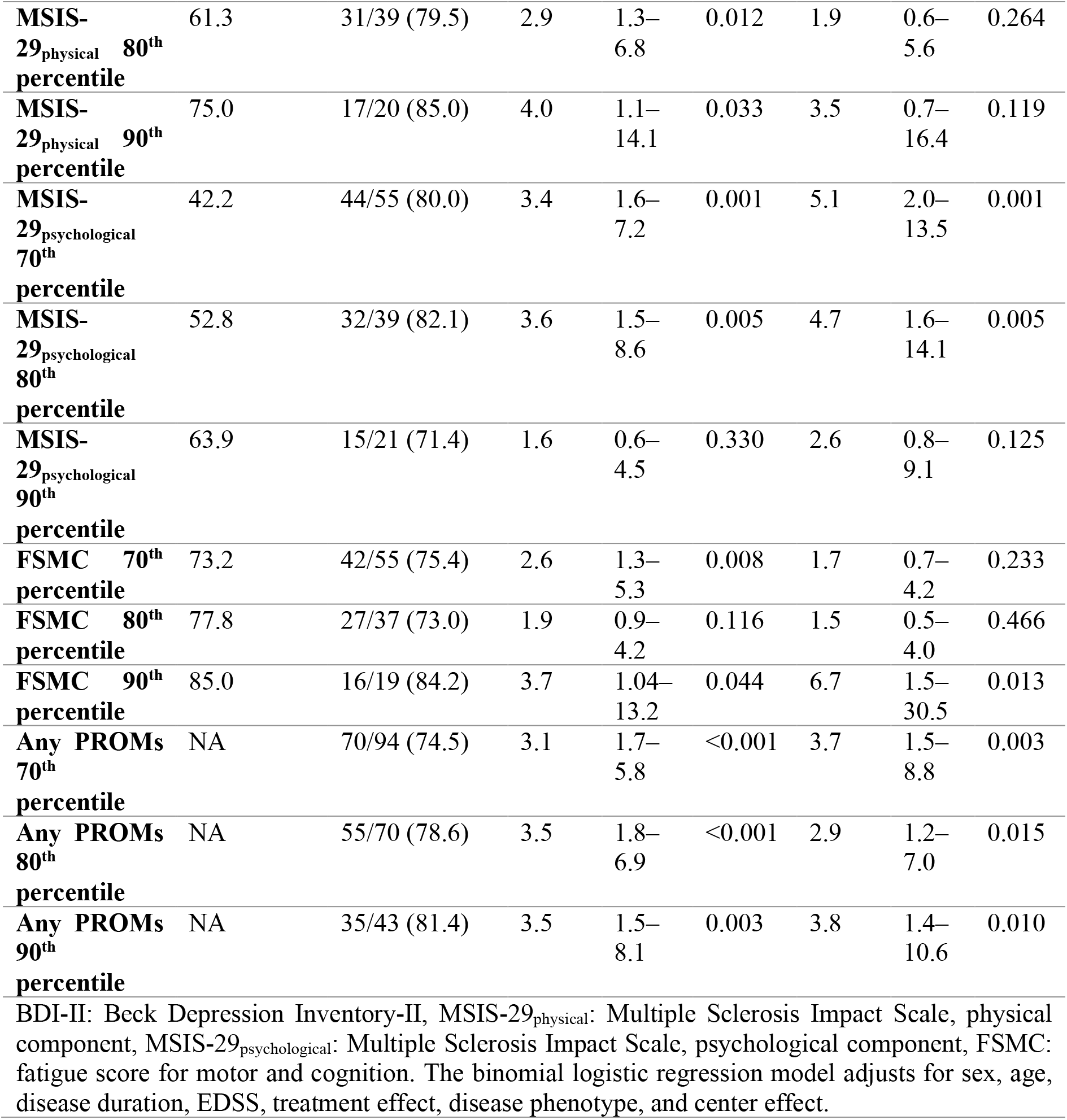
Sensitivity analysis of the various patient-reported outcome measurements (PROMs)

## Discussion

Tracking the evolution of the disease progression in PMS is challenging. In this multicentric cohort, we explored the potentials of PROMs to reflect the disability worsening over the previous two years in PMS patients. We report worse PROM scores in PMS patients with disease progression in the last two years. PROMs are increasingly implicated in the clinical care of MS patients. They offer an objective, structural way to communicate various symptoms such as fatigue and depression. Most of those symptoms are prevalent among patients with PMS [17, 18]. Besides the mobility restrictions, those symptoms significantly contribute to the reduced overall quality of life in MS patients. Our results are in line with those of a recent analysis, which suggested worsening of patient-reported outcomes (performance scales, Patient Health Questionnaire-9, European Quality of Life-5-Dimensions) in RRMS shortly before conversion into SPMS [17]. In addition, higher MSIS-29 scores were associated with reduced survival time in MS patients [19].

Worse PROMs in patients with progression can be due to the increase in the physical and psychological burden of the disease due to limited mobility with higher EDSS scores. Therefore, the included PROMs in our study might reflect the patient’s perception of the added physical disability. Nevertheless, a pathophysiological correlation cannot be excluded. Indeed, PROMs have been linked to various central nervous system structures typically impacted in PMS [20, 21]. For example, fatigue has been associated with the thalamus and cortical gray matter volumes [22-25] and, recently, brain stem pathology in MS [26]. Similarly, depression was generally correlated with atrophy parameters in temporal lobe volume and gray matter volume [27, 28].

While previous studies demonstrated improved PROM scores in patients who received various immunomodulatory treatments in RRMS [29-32], we found no difference between treated and untreated patients in terms of PROMs. This might be explained by the limited effectiveness of many DMTs in PMS, especially in older individuals, who constitute the majority of the patients in our study. Patients treated with recently approved medications like ocrelizumab represented a minority of our cohort at baseline. However, our analysis might be underpowered as the treated patients received heterogenous DMTs and constitute less than half of the included population.

The number of progression events (62%) is higher than the previous reports from the PROMISE, OLYMPUS, ORATORIO and EXPAND trials (all < 40%) [33-36] and even higher than that of our expectations during the study power calculation[10]. A possible cause might be the setting of our cohort, as only highly specialized referral MS centers were included, hinting at a potential selection bias. Moreover, the recruitment of patients began in parallel to the approval of recent medications that have demonstrated efficacy in PMS like ocrelizumab and siponimod, which might have led to more referrals of patients with suspected progression to specialized centers to initiate treatment[36]. In addition, our study population and the definition of progression differ from those of the studies mentioned above. All factors taken together could explain the higher proportion of PPMS patients and patients with clinically evident progression in our cohort compared to previous natural history studies and clinical trials in PMS.

This study has some limitations. First, the baseline progression was assessed in most cases based solely on the documented EDSS scores during the last two years, as the T25FW and 9-HPT still have not been regularly reported in routine clinical settings. Second, the PROMs are part of the secondary outcomes of our trials and were, therefore, not necessarily a part of our power calculation. Considering the overlap of single PROMs scores between the two groups (EDP vs. NEDP), longitudinal evaluation of PROMs dynamics might be more informative than a single time point. Beyond that, the utility of PROMs to predict future disability accumulation has not been addressed in this analysis. All those limitations are addressed in our current follow-up period of the study.

Taken together, our results provide evidence supporting that a more “patient-centered” approach can substantially contribute to a better classification of the patients and be subsequently considered as a valuable addition to the treatment decision algorithm. A simple, cost-efficient, and remotely accessible method to detect disability worsening could be objective and structured patient-doctor communication.

## Data Availability

The data that support the findings of this study are available from the corresponding author, Hayrettin Tumani, upon reasonable request.

## Acknowledgments

We want to thank all our patients for their engagement. We also thank our study nurses, technicians, and physicians of the participating centers for actively supporting this project. We want to thank Prof. Martin Kerschensteiner and Prof. Reinhard Hohlfeld for their support during the planning and designing of the study. Moreover, we want to thank the German MS Society, Federal Association (DMSG) for its generous funding, the German MS trust, and the AMSEL and Bavarian MS Trust for their ongoing support.

